# Favipiravir for the Treatment of Coronavirus Disease 2019; a propensity score-matched cohort study

**DOI:** 10.1101/2021.11.29.21267042

**Authors:** Rand A. Alattar, Shiema Abdalla, Tasneem A.K. Abdallah, Rashid Kazman, Aseelah Qadmour, Tawheeda B. H. Ibrahim, Bassem Alhariri, Shahd H. Shaar, Abeer Bajwa, Abeir B. Alimam, Rabia Qazi, Fatma Ben Abid, Joanne Daghfal, Ali M. Eldeeb, Kinda Shukri, Ahmed Elsayed, Fatima Rustom, Musaed Alsamawi, Alaaeldin Abdelmajid, Miguel A. P. Basulto, Armando A. R. Cobian, Mohamed Abukhattab, Muna A. Almaslamani, Abdullatif Alkhal, Ali S. Omrani

## Abstract

**Background:** We investigated clinical outcomes of favipiravir in patients with COVID-19 pneumonia.

**Methods:** Patients who between 23 May 2020 and 18 July 2020 received ≥24 hours of favipiravir were assigned to the favipiravir group, while those who did not formed the non-favipiravir group. The primary outcome was 28-day clinical improvement, defined as two-category improvement from baseline on an 8-point ordinal scale. Propensity scores (PS) for favipiravir therapy were used for 1:1 matching. Cox regression was used to examine associations with the primary endpoint.

**Results:** The unmatched cohort included 1,493 patients, of which 51.7% were in the favipiravir group, and 48.3% were not receiving supplemental oxygen at baseline. Favipiravir was started within a median of 5 days from symptoms onset. Significant baseline differences between the two unmatched groups existed, but not between the PS-matched groups (N = 774). After PS-matching, there were no significant differences between the two groups in the proportion with 28-day clinical improvement (93.3% versus 92.8%, P 0.780), or 28-day all-cause mortality (2.1% versus 3.1%, P 0.360). Favipiravir was associated with more viral clearance by day 28 (79.8% versus 64.1%, P <0.001). In the adjusted Cox proportional hazards model, favipiravir therapy was not associated 28-day clinical improvement (adjusted hazard ratio 0.978, 95% confidence interval 0.862 –1.109, P 0.726). Adverse events were common in both groups, but the 93.9% were Grades 1–3.

**Conclusion:** Favipiravir therapy for COVID-19 pneumonia is well tolerated but is not associated with an increased likelihood of clinical improvement or reduced all-cause mortality by 28 days.

## 1. Background

By the mid-June 2021, more than 175 million confirmed cases of Severe Acute Respiratory Coronavirus 2 (SARS-CoV-2), the causative agent of Coronavirus Diseases 2019 (COVID-19), had been reported globally, with approximately 3.8 million associated deaths.[1] Efforts to identify potential antivirals therapies for COVID-19 have focused largely on repurposing existing agents that had been in clinical use or in different stages of development for other indications. Hydroxychloroquine and azithromycin, each alone or in combination, and lopinavir-ritonavir (LPV/r) were found to be ineffective, while remdesivir was shown to be beneficial in COVID-19 patients who require non-invasive oxygen support.[2-5]

Favipiravir, a prodrug that is ribosylated and phosphorylated intracellularly to form the active metabolite favipiravir ibofuranosyl-5′-triphosphate (T-705-RTP), is a competitive inhibitor of RNA-dependent RNA polymerase with *in vitro* activity against influenza viruses, several haemorrhagic fever viruses, and SARS-CoV-2.[6, 7] Interest in the potential role of favipiravir for the treatment of patients with COVID-19 was initially sparked by a small observational study from China which had suggested that combining inhaled interferon-alpha with favipiravir was associated with significantly shortened time to SARS-CoV-2 clearance and with earlier radiological improvement compared with its combination with LPV/r.[8] A pilot, open-label, randomized trial of 60 patients from Russia also suggested faster viral clearance and earlier defervescence with favipiravir compared with standard care alone.[9] Several case series, observational studies of varying sizes, and underpowered randomized trials followed, but their results have been inconsistent and inconclusive.[10, 11] The aim of this study was to investigate the clinical outcomes and safety of favipiravir in a large cohort of patients hospitalised COVID-19 pneumonia.

## 2. Materials and Methods

### 2.1. Patients and materials

The study was undertaken at Hamad Medical Corporation (HMC), the provider of all COVID-19 medical care for the 2.8 million population of Qatar. SARS-CoV-2 infection was confirmed by real-time polymerase chain reaction (RT-PCR) assays TaqPath COVID-19 Combo Kit (Thermo Fisher Scientific, Waltham, Massachusetts) or Cobas SARS-CoV-2 Test (Roche Diagnostics, Rotkreuz, Switzerland) on respiratory tract specimens. We used the HMC COVID-19 database to identify patients aged 18 years or more who were hospitalised with RT-PCR-confirmed SARS-CoV-2 pneumonia during the period between 23 May 2020 and 18 July 2020. Patients who received 24 hours or more of favipiravir therapy were assigned to the favipiravir group (FVP group), while the non-favipiravir group (non-FVP group) included those who did not receive favipiravir. Favipiravir was administered orally, as two 1600 mg doses 12 hourly for one day, followed by 600 mg twice daily for up to nine more days. Hydroxychloroquine was given at a dose of 400 mg 12 hourly for one day, followed by 400 mg daily for up to 6 more days, while lopinavir (400 mg)-ritonavir (100 mg) was given 12 hourly for up to seven days. Treatment selection was according to the treating physicians at the time of the of initiation.

COVID-19 severity was categorised according to an eight-point ordinal scale.[5] The categories were: 1, not hospitalised and without limitations of activities; 2, not hospitalised but has limitation of activities, requiring oxygen, or both; 3, hospitalised, not requiring supplemental oxygen and no longer requiring ongoing medical; 4, hospitalised, not requiring supplemental oxygen but requiring ongoing medical care; 5, hospitalised and requiring any supplemental oxygen; 6, hospitalised and requiring non-invasive ventilation (NIV) or use of high-flow nasal oxygen (HFNO) devices; 7, hospitalised and receiving invasive mechanical ventilation (IMV) or extracorporeal membrane oxygenation (ECMO); and 8, death.

Clinical and laboratory data were retrieved retrospectively from the electronic healthcare system during the period from 19 July 2020 to 14 August 2020. The presence of radiological evidence of pneumonia was verified by two radiologists who were blinded to the patients’ study allocation. Adverse events were defined and graded according to the Common Terminology Criteria for Adverse Events (CTCAE), Version 5.0, 2017.

### 2.2. Outcomes

The primary outcome was clinical improvement by day 28. Clinical improvement was defined as two-category improvement from baseline on the ordinal severity scale. Secondary outcomes were all-cause mortality at 28 days, the proportion with ordinal scale score of 3 or lower at day 28, hospital length of stay, and viral clearance (defined as one SARS-CoV-2 RT-PCR with a cycle threshold of >30 on a respiratory tract sample taken ≥10 days from onset of symptoms).

### 2.3. Statistical analysis

Categorical data were summarised as numbers and percentages and compared using Pearson’s chi-squared or Fisher’s exact test, as appropriate. Continuous data were presented as medians and interquartile ranges (IQR) and compared using Wilcoxon rank-sum test. Missing baseline variables were handled by using multiple imputation with chained equations. Propensity scores for receiving favipiravir, instead of non-favipiravir therapy, were calculated using a non-parsimonious multivariate logistic regression model that included all measured potential baseline predictors for treatment. A summary of mean bias across all covariates before and after matching was displayed using histogram. The propensity scores were used as a 1:1 matching variable for favipiravir/non-favipiravir, using 0.2 calliper and without replacement (data supplement file).

Cox regression was used to examine the association of the study arm with the primary endpoint. Variables with an associated P <0.0.5 in the univariate Cox regression model were included in the multivariate analysis by forward addition and adjusted by the propensity score after excluding collinearity. All P values were two-sided with a threshold of < 0.05 for statistical significance. Statistical analyses were performed using Stata Statistical Software Release 15.1 (StataCorp LLC, College Station, Texas).

## 3. Results

### 3.1. Baseline characteristics

A total of 1,493 patients were included, of which 721 (51.7%) were in the FVP group. The majority were males (1,223, 81.9%), and the overall median age was 46 years (IQR 37– 54). Diabetes (568, 38%), and hypertension (518, 34.7%) were the most frequent co-existing medical conditions. The median body mass index was 28 kg/m^2^ (25.2–31.5). Dyspnoea was present at presentation in 707 (47.4%) individuals. At baseline, nearly half of the included patients (721, 48.3%) were not receiving supplemental oxygen, while 64 (4.3%) were on NIV, and 55 (3.7%) on IMV. Bilateral pneumonia was evident in the majority (1,258, 84.3%).

In the FVP group, the median duration from onset of symptoms to starting favipiravir therapy was 5 days (IQR 3–7), and the median duration of therapy was 7 days (IQR 6–8). Hydroxychloroquine (696, 96.5%), LPV/r (558, 77.4%), and azithromycin (716, 99.3%) were the experimental anti-SARS-CoV-2 agents used in the non-FVP group. Laboratory findings and other baseline characteristics of the unmatched cohort are summarised in Table 1.

**Table 1.** Baseline characteristics before and after propensity-score matching.

| Variable | Unmatched cohort |  |  | Propensity-score matched cohort |  |  |
| --- | --- | --- | --- | --- | --- | --- |
|  | (n = 1,493) |  |  | (n = 774) |  |  |
|  | Favipiravir group<br>(n = 772) | Non-favipiravir group<br>(n = 721) | P value | Favipiravir group<br>(n = 387) | Non-favipiravir group<br>(n = 387) | P value |
| Male sex | 624 (80.8%) | 599 (83.1%) | 0.260<sup>*</sup> | 312 (80.6%) | 312 (80.6%) | 1.000<sup>*</sup> |
| Age (years) | 48 (39.50–57) | 44 (37–54) | <0.001<sup>s</sup> | 47 (38–55) | 46 (38–57) | 0.950<sup>s</sup> |
| Nationality by WHO region |  |  | <0.001<sup>†</sup> |  |  | 0.35<sup>†</sup> |
| African Region | 6 (0.8%) | 10 (1.4%) |  | 4 (1%) | 3 (0.8%) |  |
| Eastern Mediterranean Region | 281 (36.4%) | 221 (30.7%) |  | 123 (31.8%) | 126 (32.6%) |  |
| European Region | 6 (0.8%) | 5 (0.7%) |  | 3 (0.8%) | 2 (0.5%) |  |
| Region of the Americas | 6 (0.8%) | 2 (0.3%) |  | 3 (0.8%) | 0 |  |
| South-East Asian Region | 345 (44.7%) | 398 (55.2%) |  | 192 (49.6%) | 208 (53.8%) |  |
| Western Pacific Region | 128 (16.6%) | 85 (11.8%) |  | 62 (16%) | 48 (12.4%) |  |
| Diabetes mellitus | 286 (37.1%) | 282 (39.1%) | 0.410<sup>*</sup> | 146 (37.7%) | 138 (35.7%) | 0.550<sup>*</sup> |
| Hypertension | 278 (36%) | 240 (33.3%) | 0.270<sup>*</sup> | 123 (31.8%) | 126 (32.6%) | 0.820<sup>*</sup> |
| Ischaemic heart disease | 32 (4.2%) | 28 (3.9%) | 0.800<sup>*</sup> | 15 (3.9%) | 13 (3.4%) | 0.700<sup>*</sup> |
| Chronic lung disease | 37 (4.8%) | 49 (6.8%) | 0.097<sup>*</sup> | 24 (6.2%) | 22 (5.7%) | 0.760<sup>*</sup> |
| Chronic liver disease | 5 (0.7%) | 8 (1.1%) | 0.340<sup>†</sup> | 3 (0.8%) | 3 (0.8%) | 1.000<sup>†</sup> |
| Chronic kidney disease | 31 (4%) | 54 (7.5%) | 0.004<sup>*</sup> | 20 (5.2%) | 25 (6.5%) | 0.440<sup>*</sup> |
| Cancer | 15 (1.9%) | 3 (0.4%) | 0.008<sup>†</sup> | 1 (0.3%) | 1 (0.3%) | 1.000<sup>†</sup> |
| Current or past smoker | 72 (9.3%) | 50 (6.9%) | 0.250<sup>*</sup> | 32 (8.3%) | 30 (7.8%) | 1.000<sup>*</sup> |
| Body mass index (Kg/m<sup>2</sup>) | 27.8 (25–31.2) | 28.4 (25.6–32) | 0.024<sup>§</sup> | 27.9 (24.9–31.2) | 28.4 (25.3–31.9) | 0.260<sup>§</sup> |
| Dyspnoea | 380 (49.2%) | 327 (45.4%) | 0.13<sup>*</sup> | 186 (48.1%) | 193 (49.9%) | 0.610<sup>*</sup> |
| Systolic blood pressure (mmHg) | 115 (106–126) | 108 (100–119) | <0.001<sup>§</sup> | 111 (104–121) | 112 (102–122) | 0.800<sup>§</sup> |
| Temperature (Celsius) | 38 (37.2–38.6) | 38 (37.4–38.9) | <0.001<sup>§</sup> | 38 (37.2–38.6) | 37.9 (37.2–38.7) | 0.940<sup>§</sup> |
| Heart rate (per minute) | 98 (88–110) | 97 (89–108) | 0.420<sup>§</sup> | 99 (89–110) | 98 (90–108) | 0.950<sup>§</sup> |
| Respiratory rate (per minute) | 21 (20–26) | 23 (20–28) | <0.001<sup>§</sup> | 22 (20–28) | 22 (20–26) | 0.240<sup>§</sup> |
| Oxygen saturation | 0.96 (0.94–0.97) | 0.94 (0.91–0.96) | <0.001<sup>§</sup> | 0.95 (0.92–0.97) | 0.95 (0.93–0.97) | 0.570<sup>§</sup> |
| Hydroxychloroquine therapy | 37 (4.8%) | 696 (96.5%) | <0.001<sup>*</sup> | 24 (6.2%) | 372 (96.1%) | <0.001<sup>*</sup> |
| Azithromycin therapy | 354 (45.85%) | 716 (99.3%) | <0.001<sup>*</sup> | 184 (47.6%) | 386 (99.7%) | <0.001<sup>*</sup> |
| Lopinavir/ritonavir therapy | 68 (8.8%) | 558 (77.4%) | <0.001 | 34 (8.8%) | 302 (78%) | <0.001 |
| Tocilizumab therapy | 50 (6.5%) | 99 (13.7%) | <0.001<sup>*</sup> | 31 (8%) | 27 (7%) | 0.590<sup>*</sup> |
| Systemic corticosteroids | 488 (63.2%) | 283 (39.3%) | <0.001<sup>*</sup> | 169 (43.7%) | 176 (45.5%) | 0.610<sup>*</sup> |
| Renal replacement therapy | 26 (3.4%) | 43 (6%) | 0.017<sup>*</sup> | 13 (3.4%) | 21 (5.4%) | 0.160<sup>*</sup> |
| Haemoglobin (g/dL) | 14 (12.7–15.1) | 14.1 (13.1–15) | 0.140<sup>§</sup> | 14 (12.8–15.2) | 14.2 (13.1–15) | 0.720<sup>§</sup> |
| White blood cells (x10<sup>9</sup>/L) | 6.1 (4.8–7.9) | 6.40 (5.1–8.2) | 0.070<sup>§</sup> | 6.2 (4.9–7.9) | 6.10 (4.8–7.8) | 0.710<sup>§</sup> |
| Lymphocyte count (x10<sup>9</sup>/L) | 1.2 (0.8–1.6) | 1.2 (0.9–1.6) | 0.023<sup>§</sup> | 1.2 (0.8–1.6) | 1.3 (1–1.7) | 0.200<sup>§</sup> |
| Platelets (x10<sup>9</sup>/L) | 218 (176.5–273.5) | 219 (178–282) | 0.590<sup>§</sup> | 220 (180–274) | 218 (174–286) | 0.710<sup>§</sup> |
| Serum creatinine (μmol/L) | 85 (71–99) | 84 (69–98) | 0.230<sup>§</sup> | 86 (71–99) | 82 (67–95) | 0.026<sup>§</sup> |
| Alaine transaminase (IU/L) | 32.1 (22–55) | 34 (23.4–53) | 0.073<sup>§</sup> | 33 (23–56) | 32 (22–50) | 0.68<sup>§</sup> |
| Aspartate transaminase (IU/L) | 36 (25–56) | 39 (28–58) | 0.009<sup>§</sup> | 37 (26–58) | 37 (27–53) | 0.81<sup>§</sup> |
| C-reactive protein (mg/L) | 45.9 (18.4–93.5) | 53 (22.5–110) | 0.017<sup>\$</sup> | 46.3 (20–95.2) | 45.7 (18.6–88.5) | 0.370<sup>\$</sup> |
| Ferritin (µg/L) | 580 (293–995) | 602 (300–965) | 0.740<sup>\$</sup> | 609 (310–961) | 563 (289–951) | 0.500<sup>\$</sup> |
| D-dimer (mg/L) | 0.49 (0.34–0.83) | 0.54 (0.36–1) | 0.002<sup>\$</sup> | 0.52 (0.35–0.89) | 0.53 (0.35–0.94) | 0.330<sup>\$</sup> |
| Bilateral pneumonia | 603 (78.1%) | 655 (90.9%) | <0.001<sup>*</sup> | 333 (86.1%) | 330 (85.3%) | 0.760<sup>*</sup> |
| Baseline ordinal scale category |  |  | 0.910<sup>*</sup> |  |  | 0.130<sup>*</sup> |
| Category 4 (no supplemental oxygen) | 376 (48.7%) | 345 (47.9%) |  | 201 (51.9%) | 215 (55.6%) |  |
| Category 5 (supplemental oxygen) | 336 (43.5%) | 317 (44%) |  | 153 (39.5%) | 153 (39.5%) |  |
| Category 6 (HFNO or NIV) | 34 (4.4%) | 30 (4.2%) |  | 18 (4.7%) | 7 (1.8%) |  |
| Category 7 (IMV or ECMO) | 26 (3.4%) | 29 (4%) |  | 15 (3.9%) | 12 (3.1%) |  |

Data are shown as number (%) or median (interquartile range). <sup>\*</sup>Pearson's chi-squared test, <sup>†</sup>Fisher's exact test, <sup>\$</sup>Wilcoxon rank-sum test. ECMO, extracorporeal membrane oxygenation; HFNO, high-flow nasal oxygen; IMV, invasive mechanical ventilation; NIV, non-invasive ventilation; WHO, World Health Organization

Significant baseline differences between the two unmatched groups included older age in the FVP group, and more frequent chronic kidney disease and cancer. Furthermore, patients in the FVP group had significantly higher median baseline systolic blood pressure, oxygen saturation, and oral temperature. Tocilizumab was used more frequently in the non-FVP group, whereas systemic corticosteroids use was more prevalent in the FVP arm. Other differences between the study’s two unmatched groups are shown in (Table 1). Propensity score-matching produced a cohort with 387 individuals in each group, without any significant differences in their baseline characteristics (Table 1).

### 3.2. Outcomes

In the unmatched cohort, individuals in the FVP group were more likely to achieve clinical improvement within 28 days (93.7% versus 90.9%, P 0.042), and to have an ordinal scale category 3 or lower status by day 28 (93% versus 88.1%, P 0.001). Moreover, viral clearance was more likely in the FVP group (78.5% versus 63.4%, P <0.001). However, there were no significant differences between the two groups in 28-day all-cause mortality (2.6% versus 3.3%, P 0.400), or hospital length of stay (median 9 versus 10 days, P 0.420) (Table 2).

**Table 2.** Clinical outcomes before and after propensity-score matching.

|  | Unmatched cohort<br>(n = 1,493) |  |  | Propensity-score matched cohort<br>(n = 774) |  |  |
| --- | --- | --- | --- | --- | --- | --- |
|  | Favipiravir group<br>(n = 772) | Non-favipiravir group<br>(n = 721) | P value | Favipiravir group<br>(n = 387) | Non-favipiravir group<br>(n = 387) | P value |
| Clinical improvement within 28 days | 723 (93.7%) | 655 (90.9%) | 0.042 | 361 (93.3%) | 359 (92.8%) | 0.780 |
| Days to clinical improvement | 8.50 (6–11.3) | 8 (5–12) | 0.130<sup>§</sup> | 8.5 (6–11) | 8 (5–12) | 0.072<sup>§</sup> |
| All-cause mortality at 28 days | 20 (2.6%) | 24 (3.3%) | 0.400<sup>*</sup> | 8 (2.1%) | 12 (3.1%) | 0.360<sup>*</sup> |
| Ordinal scale category ≤3 on day 28 | 718 (93%) | 635 (88.1%) | 0.001<sup>*</sup> | 360 (93%) | 352 (91%) | 0.290<sup>*</sup> |
| Hospital length of stay | 9 (6–13) | 10 (5–16) | 0.420<sup>§</sup> | 9 (6–12) | 9 (5–14.5) | 0.440<sup>§</sup> |
| Viral clearance | 606 (78.5%) | 457 (63.4%) | <0.001<sup>†</sup> | 309 (79.8%) | 248 (64.1%) | <0.001<sup>†</sup> |
| Status on day 28 |  |  | 0.014<sup>*</sup> |  |  | 0.570<sup>*</sup> |
| Died | 20 (2.6%) | 24 (3.3%) |  | 8 (2.1%) | 12 (3.1%) |  |
| Hospital floor | 19 (2.5%) | 31 (4.3%) |  | 12 (3.1%) | 11 (2.8%) |  |
| Intensive care unit | 20 (2.6%) | 35 (4.9%) |  | 9 (2.3%) | 14 (3.6%) |  |
| Discharged | 713 (92.4%) | 631 (87.5%) |  | 358 (92.5%) | 350 (90.4%) |  |

Data are shown as number (%) or median (interquartile range). <sup>\*</sup> Pearson's chi-squared test, <sup>†</sup> Fisher's exact test, <sup>§</sup> Wilcoxon rank-sum test

On the other hand, the propensity score-matched groups did not differ significantly in the proportion with clinical improvement within 28 days (93.3% versus 92.8%, P 0.780), or the secondary endpoints of the proportion with category 3 status or less on day 28 (93% versus 91%, P 0.290), 28-day all-cause mortality (2.1% versus 3.1%, P 0.360), or hospital length of stay (median 9 days versus 9 days, P 0.440). However, favipiravir was associated with a higher proportion of viral clearance by day 28 (79.8% versus 64.1%, P <0.001). Sub-analysis by baseline need for oxygen support yielded similar results (Table S1 in the data supplement file).

Several co-variants were significantly associated with clinical improvement in the unadjusted Cox proportional hazards model (Table 3). However, the adjusted model identified older age, chronic kidney disease, all levels of oxygen support requirement at baseline, tocilizumab, and corticosteroid therapy as independent predictors of clinical improvement. Receipt of favipiravir was not associated clinical improvement by day 28 (adjusted hazard ratio 0.978, 95% confidence interval 0.862 –1.109, P 0.726) (Table 3).

**Table 3.** Cox proportional hazards for clinical improvement within 28 days.

|  | Univariate analysis |  |  | Multivariate analysis |  |  |
| --- | --- | --- | --- | --- | --- | --- |
| Covariate | Hazard Ratio | 95% confidence interval | P value | Hazard Ratio | 95% confidence interval | P value |
| Favipiravir group | 1.070 | 0.962 – 1.190 | 0.210 | 0.978 | 0.862 – 1.109 | 0.726 |
| Age | 0.978 | 0.974 – 0.982 | <0.001 | 0.983 | 0.977 – 0.988 | <0.001 |
| Male sex | 0.916 | 0.799 – 1.051 | 0.210 |  |  |  |
| smoking | 0.836 | 0.685 – 1.020 | 0.077 |  |  |  |
| Diabetes mellitus | 0.657 | 0.588 – 0.734 | <0.001 | 0.917 | 0.806 – 1.042 | 0.182 |
| Hypertension | 0.636 | 0.568 – 0.713 | <0.001 | 0.955 | 0.830 – 1.099 | 0.521 |
| Ischemic heart disease | 0.709 | 0.540 – 0.931 | 0.013 | 0.914 | 0.679 – 1.229 | 0.551 |
| Chronic lung disease | 0.707 | 0.556 – 0.898 | 0.004 | 1.056 | 0.824 – 1.352 | 0.668 |
| Chronic liver disease | 0.317 | 0.151 – 0.667 | 0.002 | 0.607 | 0.287 – 1.284 | 0.192 |
| Chronic kidney disease | 0.350 | 0.266 – 0.461 | <0.001 | 0.603 | 0.450 – 0.810 | 0.001 |
| Cancer | 0.555 | 0.328 – 0.940 | 0.029 | 0.639 | 0.365 – 1.118 | 0.116 |
| Body mass index | 0.999 | 0.988 – 1.011 | 0.929 |  |  |  |
| Baseline supplemental oxygen | 0.529 | 0.474 – 0.591 | <0.001 | 0.804 | 0.703 – 0.921 | 0.002 |
| Baseline non-invasive ventilation | 0.327 | 0.244 – 0.439 | <0.001 | 0.702 | 0.507 – 0.971 | 0.032 |
| Baseline invasive ventilation | 0.207 | 0.140 – 0.304 | <0.001 | 0.452 | 0.300 – 0.681 | <0.001 |
| Baseline systolic blood pressure | 1.002 | 0.999 – 1.005 | 0.249 |  |  |  |
| Baseline lymphocyte count | 1.003 | 0.980 – 1.027 | 0.778 |  |  |  |
| Tocilizumab therapy | 0.398 | 0.327 – 0.486 | <0.001 | 0.717 | 0.576 – 0.891 | 0.003 |
| Systemic corticosteroids | 0.467 | 0.419 – 0.520 | <0.001 | 0.490 | 0.419 – 0.573 | <0.001 |

### 3.3. Adverse events

The total number of adverse events was 1,664, of which 921 (55.3%) occurred in individuals in the FVP group. Most adverse events were classified as Grades 1–3 (1,563, 93.9%). Grade 4 adverse events were experienced by 13 (1.7%) individuals in the FVP group, compared with 36 (5%) individuals in the non-FVP group (P <0.001), while there were 15 Grade 5 adverse events, all of which were in the non-FVP group (P <0.001).

The most frequently reported adverse events were alanine transaminase (ALT) increase (498, 33.4%), aspartate transaminase (AST) increase (336, 22.5%), and corrected QT interval (QTc) prolongation (162, 10.9%). ALT and AST increase were significantly more frequent in the FVP group (P <0.001 for both), whereas QTc prolongation was more common in the non-FVP group (P 0.034). Out of 162 individuals with QTc prolongation in the favipiravir FVP, 32 (45.1%) and 6 (8.5%) had received concomitant azithromycin or hydroxychloroquine, respectively. Further details on the reported adverse events are provided in Table 4, and in Tables S2, S3, and S4 in the data supplement file.

**Table 4.** Adverse events.

|  | **Entire cohort**<br>**(n = 1,493)** | **Favipiravir**<br>**group**<br>**(n = 772)** | **Non-favipiravir**<br>**group**<br>**(n = 721)** | **P value** |
| --- | --- | --- | --- | --- |
| Any Grade 1–3 AE | 822 (55.1%) | 474 (61.4%) | 348 (48.3%) | <0.001<sup>*</sup> |
| Any Grade 4 AE | 49 (3.3%) | 13 (1.7%) | 36 (5%) | <0.001<sup>*</sup> |
| Any Grade 5 AE | 19 (1.3%) | 0 | 15 (2.1%) | <0.001<sup>†</sup> |
| **Adverse events reported in ≥1% of study subjects** |  |  |  |  |
| ALT increase | 498 (33.4%) | 304 (39.4%) | 194 (26.9%) | <0.001<sup>*</sup> |
| AST increase | 336 (22.5%) | 203 (26.3%) | 133 (18.4%) | <0.001<sup>*</sup> |
| QTc prolongation | 162 (10.9%) | 71 (9.2%) | 91 (12.6%) | 0.034<sup>*</sup> |
| Serum creatinine increase | 123 (8.2%) | 58 (7.5%) | 65 (9%) | 0.290<sup>*</sup> |
| Hyperkalaemia | 78 (5.2%) | 53 (6.9%) | 25 (3.5%) | 0.003<sup>*</sup> |
| Hyperuricaemia | 74 (5.0%) | 72 (9.3%) | 2 (0.3%) | <0.001<sup>*</sup> |
| Anaemia | 38 (2.5%) | 15 (1.9%) | 23 (3.2%) | 0.130<sup>*</sup> |
| Hyponatraemia | 34 (2.3%) | 24 (3.1%) | 10 (1.4%) | 0.026<sup>*</sup> |
| ALP increase | 30 (2.0%) | 19 (2.5%) | 11 (1.5%) | 0.200<sup>*</sup> |
| Hypernatraemia | 22 (1.5%) | 11 (1.4%) | 11 (1.5%) | 0.870<sup>*</sup> |
| Hypokalaemia | 42 (2.8%) | 18 (2.3%) | 24 (3.3%) | 0.240<sup>*</sup> |
| Diarrhoea | 18 (1.2%) | 1 (0.1%) | 17 (2.4%) | <0.001<sup>†</sup> |
| Hyperglycaemia | 18 (1.2%) | 6 (0.8%) | 12 (1.7%) | 0.120<sup>*</sup> |

AE, adverse event; ALT, alanine transferase; ALP, alkaline phosphatase; AST, aspartate transferase;
QTc, corrected QT interval. <sup>\*</sup>Pearson's chi-squared test, <sup>†</sup>Fisher's exact test

## 4. Discussion

In this propensity score-matched cohort of patients hospitalised with COVID-19 pneumonia, favipiravir was associated with a significantly higher likelihood of viral clearance, without associated clinical benefits in terms of clinical improvement or all-cause mortality by 28 days. The lack of clinical benefit in our report is consistent with findings from previous studies.[11-14] On the other hand, earlier SARS-CoV-2 clearance with favipiravir had been reported in some previous studies,[8, 9, 15, 16] but not in others.[17, 18] Higher SARS-CoV-2 viral loads in the respiratory tract have been associated with an increased risk of transmission and also increased risk of severe disease and mortality.[19, 20] Earlier SARS-CoV-2 clearance may therefore seem desirable to reduce transmission and prevent disease progression. However, the detection of SARS-CoV-2 by RT-PCR does not necessarily imply the presence of viable virus, and it is not clear that pharmacological interventions that reduce SARS-CoV-2 viral load result in improved clinical outcomes.[21]

Our negative clinical results could be related to our favipiravir dosing regimen. We used a loading daily dose of 3,200 mg of favipiravir for one day, followed by a daily dose of 1,200 mg. This is consistent with the approved favipiravir dose for influenza in Japan, albeit for a total duration of five, rather than 10 days.[10] On the other hand, the recommended favipiravir dose for Ebola virus disease is a loading dose of 6,000 mg for one day, followed by a daily dose of 2400 mg for up to 10 days. [22, 23] *In vitro*, the half maximal effective concentration (EC_50_) for favipiravir against SARS-CoV-2 is 61.88 µg/mL,[24] whereas EC_50_ for influenza viruses range from 0.01 to 3.53 µg/mL,[6, 25] and for Ebola virus it is 10 µg/mL.[7, 26] Such high EC_50_ for SARS-CoV-2 may have implications for the effective favipiravir dosing required to achieve clinical benefit in patients with SARS-CoV-2 infection.[27] However, data on the safety of higher favipiravir dosing regimens in patients with COVID-19 are limited.[10, 28]

It has been suggested that antiviral agents should be administered early in the clinical course of COVID-19, before the onset of inflammatory stage of severe COVID-19.[28] in our study, the median duration from onset of symptoms to starting favipiravir therapy was 5 days (IQR 3–7). Dabbous et al randomised 96 patients with mild to moderate COVID-19 to favipiravir or chloroquine within three days after onset of symptoms and found no significant differences between the two groups in their hospital length of stay, or need for mechanical ventilation.[29] In an open-label randomized trial from Japan, patients with mild COVID-19 who received favipiravir on day 1, were not more likely to achieve viral clearance during the first six days or shorter times to defervescence, compared to those who started treatment on day 6 of the study.[17] It is therefore unlikely that the lack of clinical benefit in our study was related to the timing of treatment initiation.

In our study, favipiravir was not associated with clinical improvement by day 28, whereas older age, chronic kidney disease and severe COVID-19 requiring any degree of oxygen support, and use of systemic steroids or tocilizumab were all independently associated with a reduced likelihood of clinical improvement. Older age and co-existing chronic medical conditions are known predictors of poor clinical outcomes in our setting.[30] Systemic corticosteroids are associated with improved survival in patients with COVID-19 who require oxygen support.[31] In our study, 48.7% of patients did not require any oxygen support at baseline. Similarly, tocilizumab was associated with reduced mortality in patients with severe COVID-19 who had C-reactive protein (CRP) levels of ≥75 mg/L.[32] The median baseline CRP in our study population was 46.3 mg/L (IQR 19.2–91.4). It is therefore not surprising that these two agents were not associated with improved rates of clinical recovery in this study.

Favipiravir, at standard dosing, is known to be safe and well-tolerated.[10, 28] Adverse events were frequent in both groups in our study. However, the vast majority were mild and did not result in premature treatment discontinuation. ALT, AST and serum uric acid elevations are known common adverse events in association with favipiravir.[9, 10] Interestingly, Doi, et al reported hyperuricaemia in 84.1% in association with favipiravir, most of which uneventful and reversible.[17]

Of note, we reported QTc prolongation in 9.2% of patients in the FVP group, and 12.6% of those in the non-FVP group (P 0.034). Nearly half of those with QTc prolongation in the FVP group had received concomitant hydroxychloroquine or azithromycin. QTc prolongation is a well-recognised adverse event in association with hydroxychloroquine and azithromycin.[33] It has also been occasionally reported in favipiravir recipients, but a causal link has not been established.[12, 34] For example, in healthy Japanese volunteers, the administration of single oral doses of favipiravir 1,200 and 2,400 mg was not associated with QTc prolongation.[35] Moreover, a retrospective study from Turkey investigated QTc prolongation in 189 COVID-19 patients who received hydroxychloroquine, favipiravir, or a combination of both. Favipiravir monotherapy was associated with a median QTc change of minus three milliseconds, compared with 5 milliseconds (P 0.028) with hydroxychloroquine, and 12 milliseconds (P <0.0001) with the combination.[36] Interestingly, QTc prolongation was observed in an Italian nurse who received high dose favipiravir for the treatment of Ebola virus disease. The patient had also received levofloxacin and mefloquine, both of which can potentially cause QTc prolongation. It is therefore not clear if the prolongation is related to higher dose favipiravir.[37]

In this study, almost all patients in the comparator arm and nearly half of those in the favipiravir group had received investigational agents including hydroxychloroquine, azithromycin, and LPV/r. The standard of care for COVID-19 evolved as emerging evidence demonstrated efficacy or futility of various interventions. In particular, the potential role of hydroxychloroquine, with or without azithromycin, has been the subject of intense debate.[38, 39] Nevertheless, most international guidelines do not support the use of either agent, alone or in combination, for COVID-19 of any degree of severity.[40-44] While the presence of those agents in the study may have contributed to the observed adverse events, it is unlikely that they influenced the assessment of favipiravir’s clinical efficacy.

To the best of our knowledge, this is the largest reported study to investigate the role of favipiravir in the treatment of patients with COVID-19 pneumonia, and the first to examine outcomes after 28 days of follow up. Nevertheless, our findings are limited by the retrospective nature of the investigation. We used propensity score matching to reduce treatment allocation bias, and multivariate Cox proportional hazards to investigate the relationship between favipiravir and the study outcomes. However, we cannot rule out residual confounding.

## 5. Conclusion

Early favipiravir therapy for patients hospitalised with COVID-19 pneumonia is associated with increased viral clearance, but not clinical improvement or all-cause mortality by 28 days. Adequately powered randomised trials are required to confirm our findings.

## Data Availability

All data produced in the present study are available upon reasonable request to the corresponding author

## Abbreviations

ALT: alanine transferase
ALP: alkaline phosphatase
AST: aspartate transferase
COVID-19: coronavirus disease 2019
CRP: C-reactive protein
CTCAE: Common Terminology Criteria for Adverse Events
ECMO: extracorporeal membrane oxygenation
EC_50_: half maximal effective concentration
HFNO: high-flow nasal oxygen
HMC: Hamad Medical Corporation
IMV: invasive mechanical ventilation
IQR: interquartile ranges
LPV/r: lopinavir-ritonavir
NIV: non-invasive ventilation
QTc: corrected QT interval
RT-PCR: real-time polymerase chain reaction
SARS-CoV-2: severe acute respiratory syndrome coronavirus

## Acknowledgements

We would like to thank all colleagues in Hamad Medical Corporation for their outstanding service and dedication.

## Data availability statement

The datasets used and analysed during the current study are available from the corresponding author on reasonable request.

## Ethics approval

The study was approved by the Institutional Review Board at Hamad Medical Corporation (MRC-01-20-994) with a waiver of informed consent.

## Authors’ contribution

Conceptualization and study design: RAA and ASO. Data curation: SA, TAKA, RK, AQ, TBHI, BA, SHS, AB, ABA, RQ, FA, AME, KS, AE, MA, and AA. Formal analysis and interpretation: JD and ASO. Resources: FR, MAPB, AARC, MA, and MAA. Writing – original draft preparation: RAA and ASO. All authors approved the version submitted for publication.

