## supplement file for "Favipiravir for the Treatment of Coronavirus Disease 2019; a propensity score-matched cohort study"

**Propensity score matching**

Propensity scores for receiving favipiravir, instead of non-favipiravir therapy, were used to balance the differences in baseline variables between the treatment groups. A non-parsimonious multivariate logistic regression model was used to estimate each individual’s propensity score. The model included the following baseline variables: sex, age, nationality by WHO region, diabetes mellitus, hypertension, ischaemic heart disease, chronic lung disease, chronic liver disease, chronic kidney disease, cancer, current or past smoker, body mass index, dyspnoea, systolic blood pressure, temperature, heart rate, respiratory rate, oxygen saturation, tocilizumab therapy, systemic corticosteroids, lymphocyte count, serum creatinine, c-reactive protein, serum ferritin, and bilateral pneumonia.

The propensity scores were used as a 1:1 matching variable for favipiravir/non-favipiravir, using 0.2 calliper and without replacement. A summary of mean bias across all covariates before and after matching was displayed using histogram (Figure S1 and S2).

**Figure S1. Standardised bias across variables before and after propensity score-matching.**

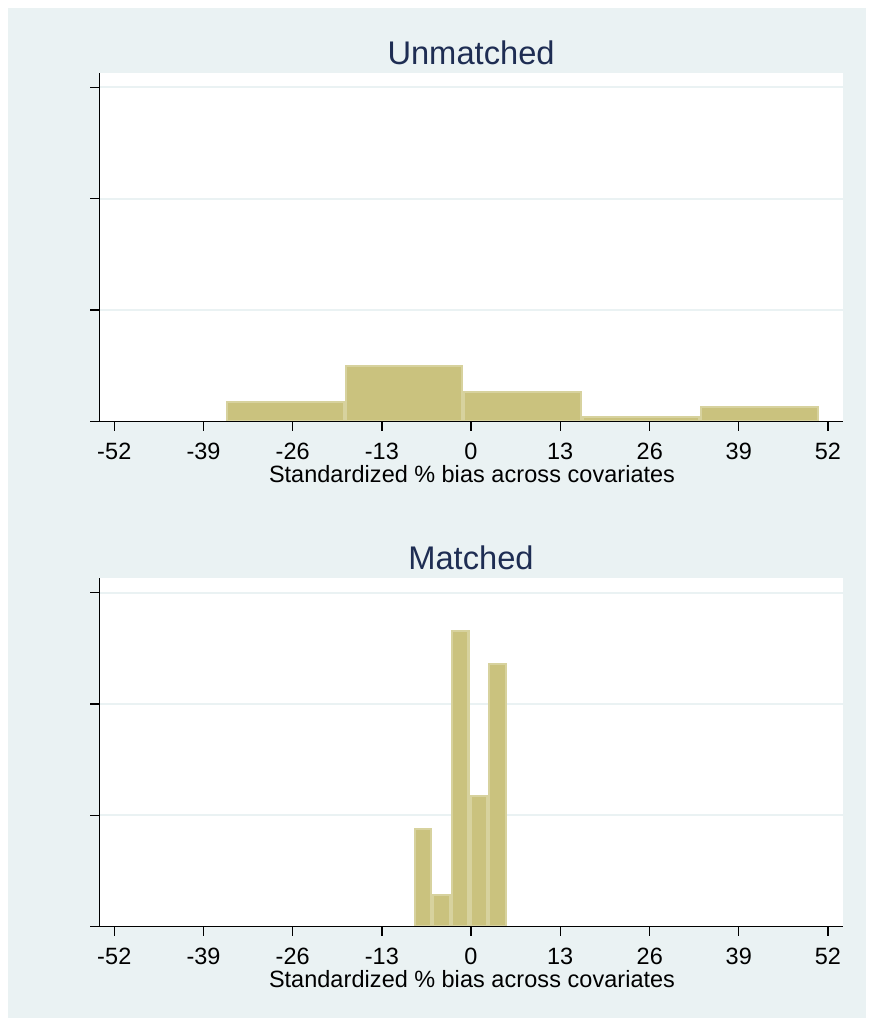

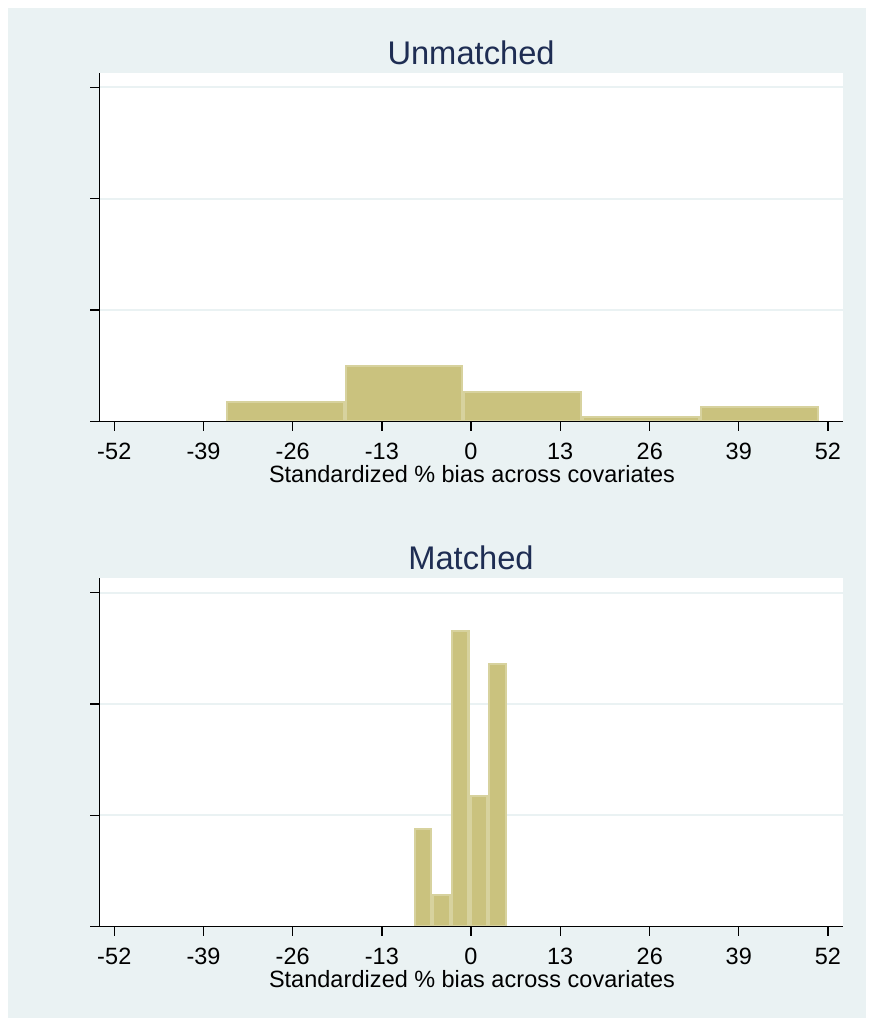

**Figure S2. Standardised differences across variables used to generate the propensity scores before and after propensity score-matching.**

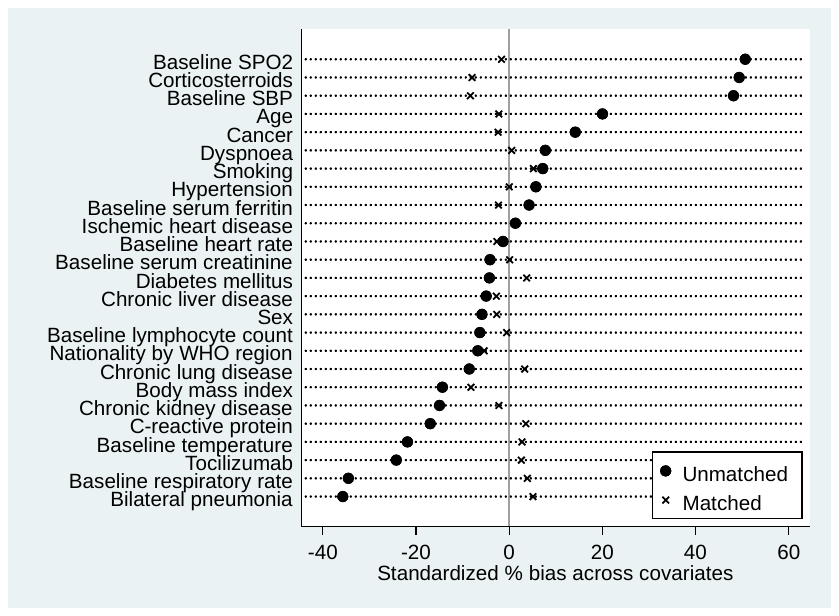

SBP, systolic blood pressure; SpO2, oxygen saturation; WHO, World Health Organization

**Table S1. Clinical outcomes by baseline oxygen requirements**

**A. Patients not requiring oxygen support at baseline**

|  | **Entire cohort** | | | **Propensity score-matched cohort** | | |
| --- | --- | --- | --- | --- | --- | --- |
|  | **Favipiravir group**  **(n = 376)** | **Non-favipiravir group (N = 345)** | **P value** | **Favipiravir group**  **(n = 201)** | **Non-favipiravir group**  **(n = 215)** | **P value** |
| **Clinical improvement within 28 days** | 370 (98.40%) | 337 (97.68%) | 0.48 | 197 (98.01%) | 209 (97.21%) | 0.59 |
| **All-cause mortality at 28 days** | 0 ( 0.00%) | 3 ( 0.87%) | 0.07 | 0 ( 0.00%) | 2 ( 0.93%) | 0.17 |
| **Ordinal scale category ≤3 on day 28** | 369 (98.14%) | 333 (96.52%) | 0.18 | 197 (98.01%) | 208 (96.74%) | 0.42 |

**B. Patients requiring oxygen support at baseline**

|  | **Entire cohort** | | | **Propensity score-matched cohort** | | |
| --- | --- | --- | --- | --- | --- | --- |
|  | **Favipiravir group**  **(n = 396)** | **Non-favipiravir group**  **(n = 376)** | **P value** | **Favipiravir group**  **(n = 186)** | **Non-favipiravir group**  **(n = 172)** | **P value** |
| **Clinical improvement within 28 days** | 353 (89.14%) | 318 (84.57%) | 0.06 | 164 (88.17%) | 150 (87.21%) | 0.78 |
| **All-cause mortality at 28 days** | 20 ( 5.05%) | 21 ( 5.59%) | 0.74 | 8 ( 4.30%) | 10 ( 5.81%) | 0.51 |
| **Ordinal scale category ≤3 on day 28** | 349 (88.13%) | 302 (80.32%) | 0.003 | 163 (87.63%) | 144 (83.72%) | 0.29 |

**Table S2. Grade 5 adverse events**

|  | **Entire cohort**  **(n = 1,493)** | **Favipiravir group**  **(n = 772)** | **Non-favipiravir group**  **(n = 721)** |
| --- | --- | --- | --- |
| Asystole | 5 | 0 | 5 |
| Creatinine increase | 3 | 0 | 3 |
| Acute coronary syndrome | 2 | 0 | 2 |
| Acute respiratory distress syndrome | 2 | 0 | 2 |
| Cardiac arrest | 2 | 0 | 2 |
| Intracranial hemorrhage | 2 | 0 | 2 |
| Multi-organ failure | 2 | 0 | 2 |
| AST increase | 1 | 0 | 1 |
| ALP increase | 1 | 0 | 1 |
| Hyperkalemia | 1 | 0 | 1 |
| Pneumothorax | 1 | 0 | 1 |
| Sepsis | 1 | 0 | 1 |
| Thromboembolic event | 2 | 0 | 2 |
| **Grand Total** | **25** | **0** | **25** |

**Table S3. Grade 4 adverse events**

|  | **Entire cohort**  **(n = 1,493)** | **Favipiravir group**  **(n = 772)** | **Non-favipiravir group**  **(n = 721)** |
| --- | --- | --- | --- |
| Creatinine increase | 23 | 7 | 16 |
| ALT increase | 12 | 2 | 10 |
| AST increase | 6 | 2 | 4 |
| Hyponatremia | 4 | 2 | 2 |
| Acute respiratory distress syndrome | 4 | 0 | 4 |
| Platelet count decrease | 4 | 0 | 4 |
| Heart failure | 3 | 0 | 3 |
| Sepsis | 3 | 0 | 3 |
| Anemia | 2 | 1 | 1 |
| Hypernatremia | 2 | 0 | 2 |
| Multi-organ failure | 2 | 0 | 2 |
| Rhabdomyolysis | 2 | 0 | 2 |
| Tachycardia | 1 | 1 | 0 |
| Cardiac conduction disorder | 1 | 0 | 1 |
| Acidosis | 1 | 0 | 1 |
| Hyperbilirubinemia | 1 | 1 | 0 |
| Hypokalemia | 1 | 0 | 1 |
| Lipase increase | 1 | 0 | 1 |
| Lymphocyte count decrease | 1 | 1 | 0 |
| Disseminated intravascular coagulation | 1 | 0 | 1 |
| Encephalopathy | 1 | 0 | 1 |
| **Grand Total** | **76** | **17** | **59** |

**Table S4. Grade 1–3 adverse events**

|  | **Entire cohort**  **(n = 1,493)** | **Favipiravir group**  **(n = 772)** | **Non-favipiravir group**  **(n = 721)** |
| --- | --- | --- | --- |
| ALT increase | 489 | 305 | 184 |
| AST increase | 330 | 202 | 128 |
| Corrected QTc interval prolongation | 163 | 72 | 91 |
| Creatinine increase | 97 | 51 | 46 |
| Hyperkalemia | 77 | 53 | 24 |
| Hyperuricemia | 74 | 72 | 2 |
| Anemia | 36 | 14 | 22 |
| Hyponatremia | 30 | 22 | 8 |
| ALP increase | 29 | 19 | 10 |
| Hypernatremia | 20 | 11 | 9 |
| Hypokalemia | 20 | 2 | 18 |
| Diarrhea | 18 | 1 | 17 |
| Hyperglycemia | 18 | 6 | 12 |
| Hemolysis | 12 | 1 | 11 |
| Cardiac troponin T increase | 10 | 5 | 5 |
| CPK increase | 10 | 6 | 4 |
| Lymphocyte count decrease | 9 | 9 | 0 |
| Vomiting | 9 | 0 | 9 |
| Platelet count decrease | 8 | 1 | 7 |
| Bradycardia | 7 | 5 | 2 |
| Acute coronary syndrome | 6 | 2 | 4 |
| Heart failure | 6 | 4 | 2 |
| Hypoalbuminemia | 6 | 0 | 6 |
| Platelet count increase | 5 | 2 | 3 |
| Neutrophil count decrease | 5 | 5 | 0 |
| Hypertension | 4 | 2 | 2 |
| Hypertriglyceridemia | 4 | 0 | 4 |
| LDH increase | 4 | 3 | 1 |
| Gastrointestinal bleeding | 4 | 2 | 2 |
| Hypophosphatemia | 3 | 3 | 0 |
| Sepsis | 3 | 0 | 3 |
| Cardiac conduction disorder | 3 | 2 | 1 |
| Tachycardia | 3 | 3 | 0 |
| Constipation | 3 | 2 | 1 |
| Hyperbilirubinemia | 3 | 1 | 2 |
| Nausea | 3 | 1 | 2 |
| Hypercalcemia | 2 | 0 | 2 |
| Hypermagnesemia | 2 | 2 | 0 |
| Rhabdomyolysis | 2 | 0 | 2 |
| Abdominal pain | 2 | 1 | 1 |
| Acidosis | 2 | 0 | 2 |
| Diabetic ketoacidosis | 2 | 1 | 1 |
| WBC increase | 2 | 2 | 0 |
| WBC decrease | 1 | 0 | 1 |
| Blood bicarbonate decrease | 1 | 1 | 0 |
| Stroke | 1 | 0 | 1 |
| Delirium | 1 | 0 | 1 |
| Disseminated intravascular coagulation | 1 | 0 | 1 |
| Encephalopathy | 1 | 0 | 1 |
| Fever | 1 | 0 | 1 |
| Gastritis | 1 | 1 | 0 |
| Hyperuricemia | 1 | 1 | 0 |
| Hypoglycemia | 1 | 0 | 1 |
| Myocarditis | 1 | 1 | 0 |
| Pericarditis | 1 | 1 | 0 |
| Osteomyelitis | 1 | 1 | 0 |
| Palpitations | 1 | 1 | 0 |
| Pancreatitis | 1 | 0 | 1 |
| T wave abnormality | 1 | 1 | 0 |
| Thromboembolic event | 1 | 0 | 1 |
| Tracheoesophageal fistula | 1 | 1 | 0 |
| **Grand Total** | **1563** | **904** | **659** |
